# SARS-CoV-2 surveillance in Italy through phylogenomic inferences based on Hamming distances derived from functional annotations of SNPs, MNPs and InDels

**DOI:** 10.1101/2021.05.25.21257370

**Authors:** Adriano Di Pasquale, Nicolas Radomski, Iolanda Mangone, Paolo Calistri, Alessio Lorusso, Cesare Cammà

## Abstract

**Background:** Faced to the ongoing global pandemic of coronavirus disease, the ‘National Reference Centre for Whole Genome Sequencing of microbial pathogens: database and bioinformatic analysis’ (GENPAT) formally established at the ‘Istituto Zooprofilattico Sperimentale dell’Abruzzo e del Molise’ (IZSAM) in Teramo (Italy) supports the genomic surveillance of the SARS-CoV-2. In a context of SARS-CoV-2 surveillance needed proper and fast assessment of epidemiological clusters from large amount of samples, the present manuscript proposes a workflow for identifying accurately the PANGOLIN lineages of SARS-CoV-2 samples and building of discriminant minimum spanning trees (MST) bypassing the usual time consuming phylogenomic inferences based on multiple sequence alignment (MSA) and substitution model.

**Results:** GENPAT constituted two collections of SARS-CoV-2 samples. The samples of the first collection were isolated by IZSAM in the Abruzzo region (Italy), then shotgun sequenced and analyzed in GENPAT (n = 1 592), while those of the second collection were isolated from several Italian provinces and retrieved from the reference Global Initiative on Sharing All Influenza Data (GISAID) (n = 17 201). The main outcomes of the present study showed that (i) GENPAT and GISAID identified identical PANGOLIN lineages, (ii) the PANGOLIN lineages B.1.177 (i.e. historical in Italy) and B.1.1.7 (i.e. ‘UK variant’) are major concerns today in several Italian provinces, and the new MST-based method (iii) clusters most of the PANGOLIN lineages together, (iv) with a higher dicriminatory power than PANGOLIN, (v) and faster that the usual phylogenomic methods based on MSA and substitution model.

**Conclusions:** The shotgun sequencing efforts of Italian provinces, combined to a structured national system of metagenomics data management, provided support for surveillance SARS-CoV-2 in Italy. We recommend to infer phylogenomic relationships of SARS-CoV-2 variants through an accurate, discriminant and fast MST-based method bypassing the usual time consuming steps related to MSA and substitution model-based phylogenomic inference.

## INTRODUCTION

The coronavirus disease 19 (COVID-19) responsible of the current pandemic is due to a novel coronavirus (CoV) named SARS-CoV-2 [1]. COVID-19 was first reported in December 2019 in the city of Wuhan, Hubei Province, China, in humans largely connected to the Huanan seafood wholesale market where different species of farm and wild animals are commonly sold, although the role of the market in virus emergence is still uncertain [2–4]. To date (May 2021), 222 countries were impacted by the SARS-CoV-2 with 153 527 666 coronavirus cases, 3 217 267 deaths, 130 881 394 of recovered people, 680 364 daily new cases, and 9 981 daily deaths [5]. With more than 1 000 cases confirmed till the 1^st^ March 2020, Italy was one of the first European countries to face the SARS-CoV-2 burden [6]. At the national level, the Italian Civil Protection Department counted today 4 044 762 total cases, 121 177 deaths, 3 492 679 recovered people and 430 906 active cases in Italy [7]. COVID-19 is mainly a respiratory infection, with the most common symptoms comprising fever, dry cough, and shortness of breath [8]. About 20% of infected patients may develop severe disease, and a small percentage (5%) may become critically ill [8]. Patients with severe COVID-19 disease usually develop pneumonia or acute respiratory distress syndrome (ARDS), a condition that may require mechanical ventilation and intensive care unit treatment [8]. ARDS is often fatal [9].

CoVs have enveloped single-stranded RNA genomes with exceptional genetic complexity and variety [10]. One major contributing factor to CoV diversity is high-frequency RNA recombination [2, 3]. New sero- and biotypes have arisen from homologous RNA recombination, while heterogeneous RNA recombination events with non-coronaviral donor RNAs have led to the acquisition of novel genes [11]. SARS-CoV-2 is paradigmatic of these evolutionary mechanisms as it emerged through recombination of SARS-related coronaviruses (SARSr-CoVs) [2, 3, 12]. In addition to recombination events, the point mutations induced by replication errors play also a major role in the SARS-CoV-2 evolution (i.e. single nucleotide polymorphisms (SNPs), multi-nucleotide polymorphisms (MNPs) and small insertions/deletions (InDels)) [13]. The theoretical SNP-based mutation rate of the SARS-CoV-2 (∼10^−6^ nt^-1^ cycle^-1^) is considered low in comparison to influenza (∼3 × 10^−5^ nt^-1^ cycle^-1^) or to other RNA viruses mutating at high rates [14]. Indeed, the SARS-CoV-2 has the capacity to repair part of duplication errors caused by the RNA-dependent RNA polymerases (RdRp) [15]. Nevertheless, a SARS-CoV-2 population in one milliliter of sputum (i.e. around 10^7^ RNAs) with this theoretical mutation rate (∼10^−6^ nt^-1^ cycle^-1^) would have a high potential of evolution harboring more than one mutation in every nucleotide [16], not including the fact that the circulation over millions of individuals promotes accumulation of mutations in relatively short time.

In addition to negative impacts of the SARS-CoV-2 on hospital workload [17], medical clinic organization [18], long-term health [19], small business [20], socio-economic system [21] and employment [22], the national health care systems have to face the need for thousands of laboratory tests per day [23]. Under the authority of the Italian Ministry of Health, the Veterinary Public Health Institutes, namely Istituti Zooprofilattici Sperimentali (IZS), are involved in the diagnosis of SARS-CoV-2 through testing nasopharyngeal swabs by RT-PCR [23]. In the context of this pandemic crisis, the “National Reference Centre for Whole Genome Sequencing of microbial pathogens: database and bioinformatic analysis” (GENPAT) formally established at the IZS dell’Abruzzo e del Molise (IZSAM) in Teramo (G.U.R. 196, August 23, 2017), focuses its activities on the improvement of the analytical workflows of SARS-CoV-2 sequences produced during routine surveillance activities.

Different international teams proposed analytical workflows to reconstruct SARS-CoV-2 genomes based on *de novo* assemblies (e.g. coronaSPAdes [24], Trinity [25]) and/or consensus sequences (e.g. iVar [26]) from variant calling analysis (SAMtools [27], Freebayes [28], genomic analysis toolkit (GATK4) [29]) performed through mapping of reads (e.g. Minimap [27], Minimap2 [28], BWA [29, 30], Bowtie2 [25]) against the reference genome Wuhan-Hu-1/2019. At the international level, the resulted *de novo* assemblies and consensus sequences are commonly submitted to the Global Initiative on Sharing All Influenza Data (GISAID). Then, these *de novo* assemblies and consensus sequences are usually aligned between each other through multiple sequence alignment (MSA) (e.g. Muscle [27], Augur toolkit [31], MAFFT [32], Clustal (O/W) [33]) in order to perform substitution model-based phylogenomic inferences through maximum likelihood (ML) (e.g. IQ-TREE [31], RaxML [27]) or Bayesian models (BEAST [33]). The aligned *de novo* assemblies and consensus sequences can also be derived (i.e. snp-sites) into variant calling format (i.e. VCF) [33]. Because the biological effects of variants (i.e. SNPs, MNPs and InDels, so-called genotypes in VCF files) are required for identifying SARS-CoV-2 lineages (e.g. PANGOLIN [34]) and accordingly designing of vaccines [35], these VCF files from variant calling or aligned sequences are usually used as input of functional annotation of variants with SNPeff [27, 33] or ANNOVAR [28]. While *de novo* assembly (i.e. coronaSPAdes [24], Trinity [36]), mapping of reads (i.e. Minimap [37], Minimap2 [38], BWA [39], Bowtie2 [40]) and variant calling analysis (i.e. SAMtools [41], Freebayes [42], GATK4 [43]) are pretty fast, these SARS-CoV-2 workflows are today mainly limited by the time consuming steps aiming at performing MSA (i.e. Augur toolkit [44], MAFFT [45], Muscle [46], Clustal W [47], Clustal O [48], CMSA [49]), then substitution model-based phylogenomic inferences (i.e. IQ-TREE [50], RaxML [51], BEAST [52]). Indeed, the phylogenetic inferences based on MSA and substitution model can last from several days to several weeks depending of the computing power of facilities, especially when the collection of samples includes several hundreds of genomes.

In the area of surveillance dedicated to bacteria including *Enterococcus* [53], *Mycoplasma* [54], *Pseudomonas* [55], *Mycobacterium* [56], *Brucella* [57] and many others, coregenome multi-locus sequence typing (cgMLST) and corresponding schemes of alleles have been proposed to identify epidemiological relationships based on screening of alleles through several hundred or thousands of orthologous genes, so-called loci [58]. In comparison with the so-called allele scheme, the combination of these MLST allele numbers from a single strain allows assignation of a MLST sequence type (ST) already shared between laboratories or a new one by default [59]. The output of cgMLST methods from different analytical workflows (e.g. chewBBACA [60], SeqSphere^+^ [61], MLSTar [62], BIGSdb-Pasteur [63], Bionumerics [64]) are frequently used as input for a recent minimum spanning tree (MST) algorithm (“MSTree V2”) implemented in the workflow GrapeTree in order to visualize coregenome relationships among hundreds of thousands bacterial genomes [65]. Compared to the good practices aiming at building a phylogenomic tree based on MSA, a substitution model (i.e. JC69, K80, K81, F81, HKY85, T92, TN93, or GTR) and an inference method (i.e. ML or Bayesian models) [66, 67], the construction of a MST with “MSTree V2” is theoretically faster because it implements a directional measure based on normalized asymmetric Hamming-like distances between pairs of STs assuming that one of the pair of STs is the ancestor of the other [65].

Considering the need for a accurate, discriminant and fast assessment of SARS-CoV-2 epidemiological clusters from large amount of samples, the present manuscript proposes a variant calling analysis-based workflow to accurately identify the PANGOLIN lineages of SARS-CoV-2 samples in Italy and fastly build highly discriminant MST bypassing the usual time consuming phylogenomic inferences based on multiple sequence alignment (MSA) and substitution model. More precisly, the objectives of the present manuscript are to assess (i) the GENPAT ability to identify PANGOLIN lineages compared to the reference GISAID, (ii) the diversity of PANGOLIN lineages circulating in the Italian provinces, as well as (iii) the clustering, (iv) dicriminatory power and (v) speed of the new method aiming at performing MST-based phylogenomic inference.

## RESULTS

The objectives described above (i.e. I, ii, iii, iv and v) were assessed based on two collections of SARS-CoV-2 samples from GENPAT (Additional file 1) and GISAID (Additional file 2), as well as a workflow aiming at combining variant calling analysis with the identification of PANGOLIN lineages (Figure 1) and the MST-based phylogenomic inference through an in-house algorithm called “vcf2mst.pl” (Figures 1 and 2).

**Figure 1:**
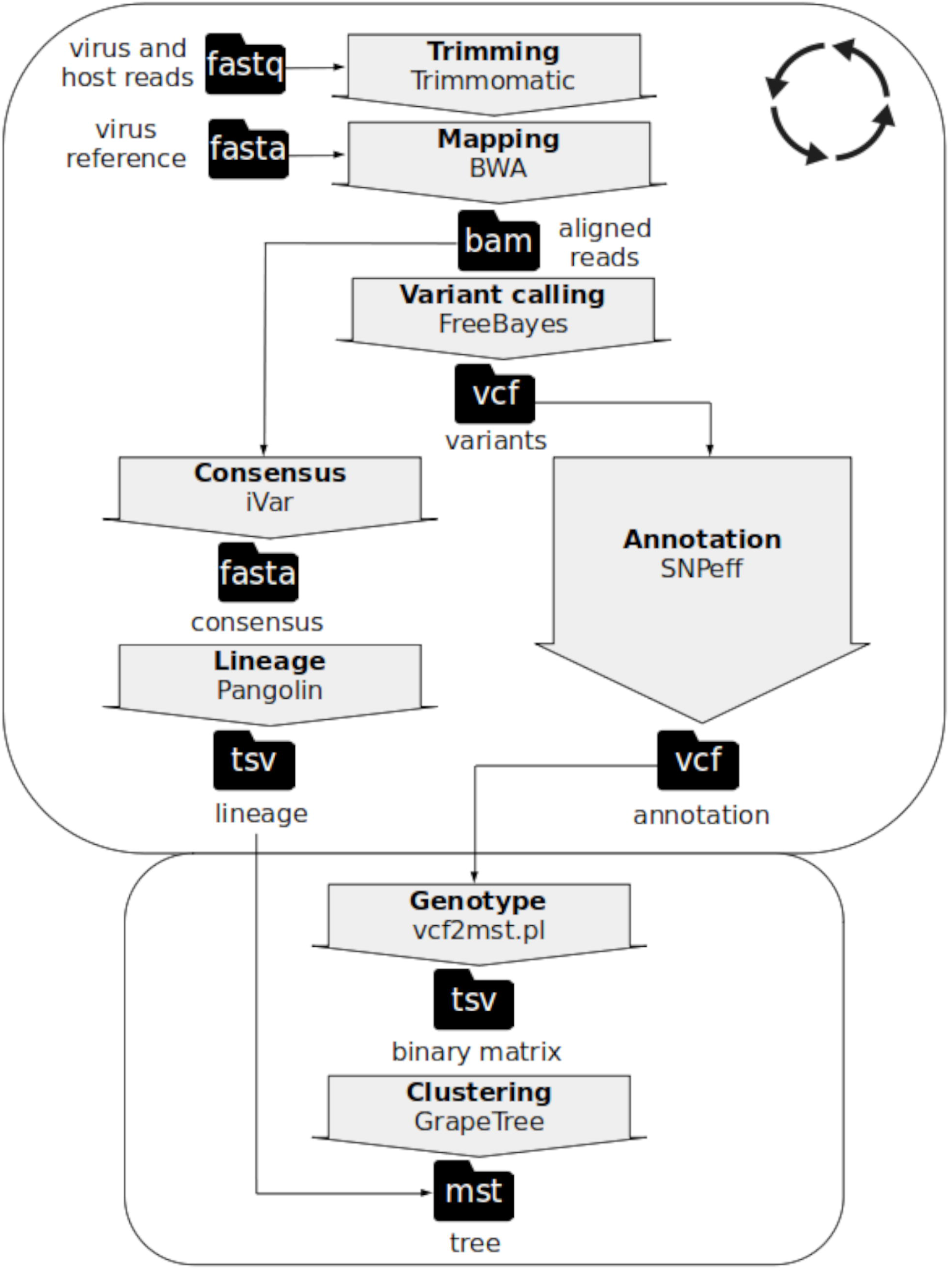
Sample dependent (rounded rectangle with circle of arrows) and dataset dependent (rounded rectangle without circle of arrows) steps of the workflow implemented in GENPAT to identify lineages of SARS-CoV-2 and build phylogenomic inference based on shotgun metagenomics paired-end read sequencing.

**Figure 2:**
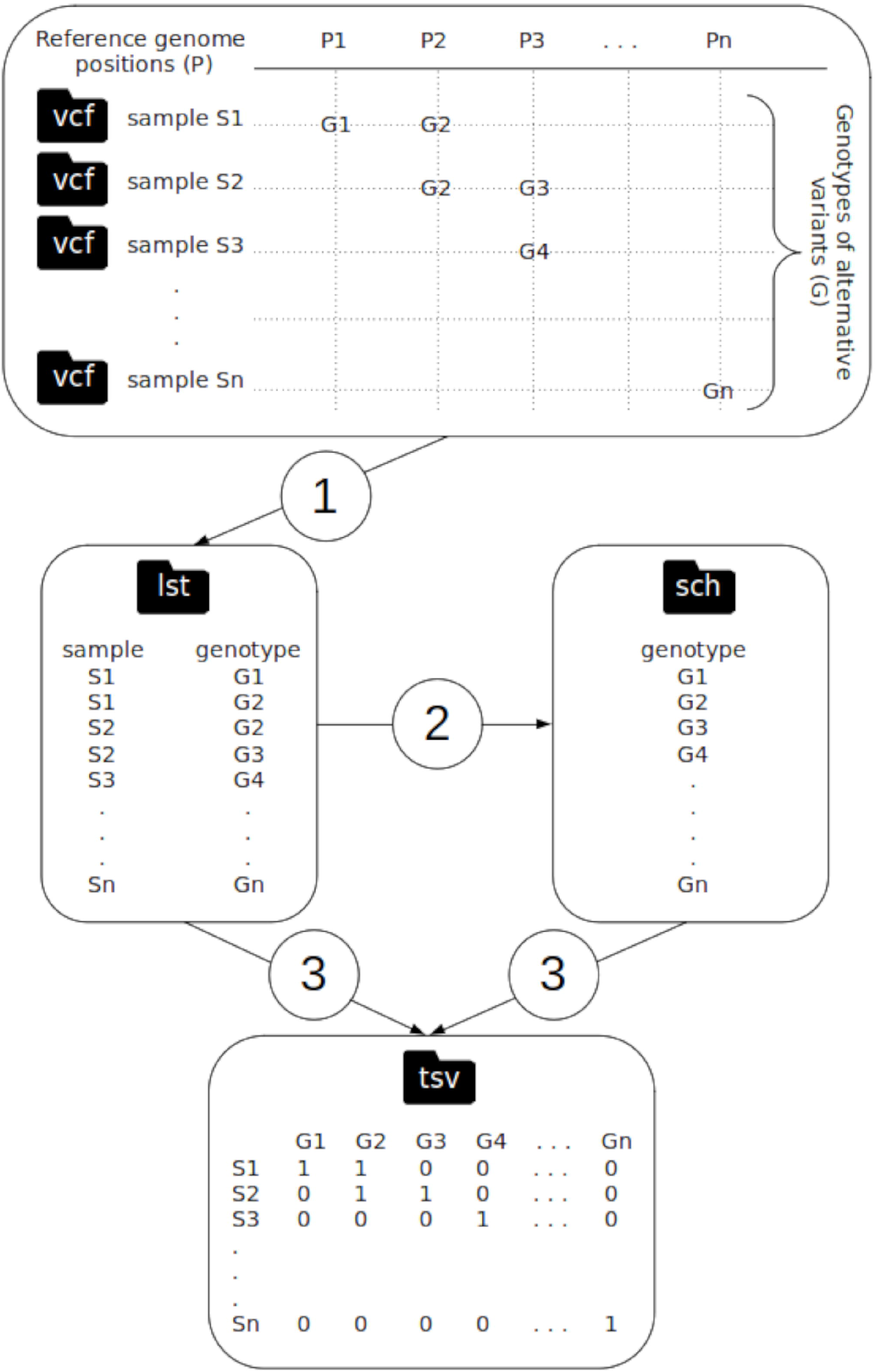
Algorithmic steps of the program “vcf2mst.pl” aiming to (1) derive functional annotations of variants (i.e. genotypes) encoded in variant calling format (vcf) into lists of samples and genotypes (lst), (2) build a scheme of genotypes (sch) and (3) create a binary matrix of genotypes according to samples involved in reference genome based-variant calling analysis (tsv) for downstream inference of minimum spanning tree (MST).

### i/ GENPAT ability to identify PANGOLIN lineages in comparison to the reference GISAID

The comparison of PANGOLIN lineages identified across the 1 592 samples from GENPAT (Additional file 1) and the 17 201 samples from GISAID (Additional file 2) allowed to estimate GENPAT capacity to identify PANGOLIN lineages in comparison to the reference GISAID. In total, 1 550 common samples between the collections GENPAT and GISAID presented identical PANGOLIN lineages (Additional files 1 and 2). These observations emphasized the accuracy of SARS-CoV-2 lineage identification implemented in the GENPAT workflow (Figure 1).

### ii/ Diversity of PANGOLIN lineages in Italy revealed by shotgun sequencing efforts of provinces

Among the SARS-CoV-2 samples from the collection GENPAT (Additional file 1, n = 1 592), the PANGOLIN lineages B.1.1.7 (62%) and B.1.177 (19%) were the mostly identified in the Abruzzo region (Figure 4C), more precisly in the provinces Chieti (32% and 5%), L’Aquila (11% and 5%), Pescara (2% and 4‰) and Teramo (16% and 8%) (Table 1 and Figure 3D). In respect of the GISAID collection (Additional file 2, n = 17 201), pics of the main PANGOLIN lineages were observed during march 2021 in Italy (Figure 3A), several Italian provinces (Figure 3B), Abruzzo region (Figure 3C) and provinces of Abruzzo region (Figure 3D). In this GISAID collection (Additional file 2), the PANGOLIN lineages B.1.1.7 (39%) and B.1.177 (17%) were also the mostly identified lineages in Italy, especially because of the province Napoli (24% and 12%) which provided a high effort of shotgun sequencing (n = 4 184 and n = 2 073) (Figure 3A and Additional file 3). In addition to Napoli province, these PANGOLIN lineages were also mainly identified in the provinces Venezia, Chieti, Bari, Trento and Teroma (Figure 3B). While the Napoli province harbored the highest effort of SARS-CoV-2 lineage identification in Italy (Figure 3D, n = 10 372: 67%), the Chieti province presented the highest effort of SARS-CoV-2 lineage identification through shotgun sequencing in the Abruzzo region (Figure 3B, n = 710: 45%).

**Table 1:**
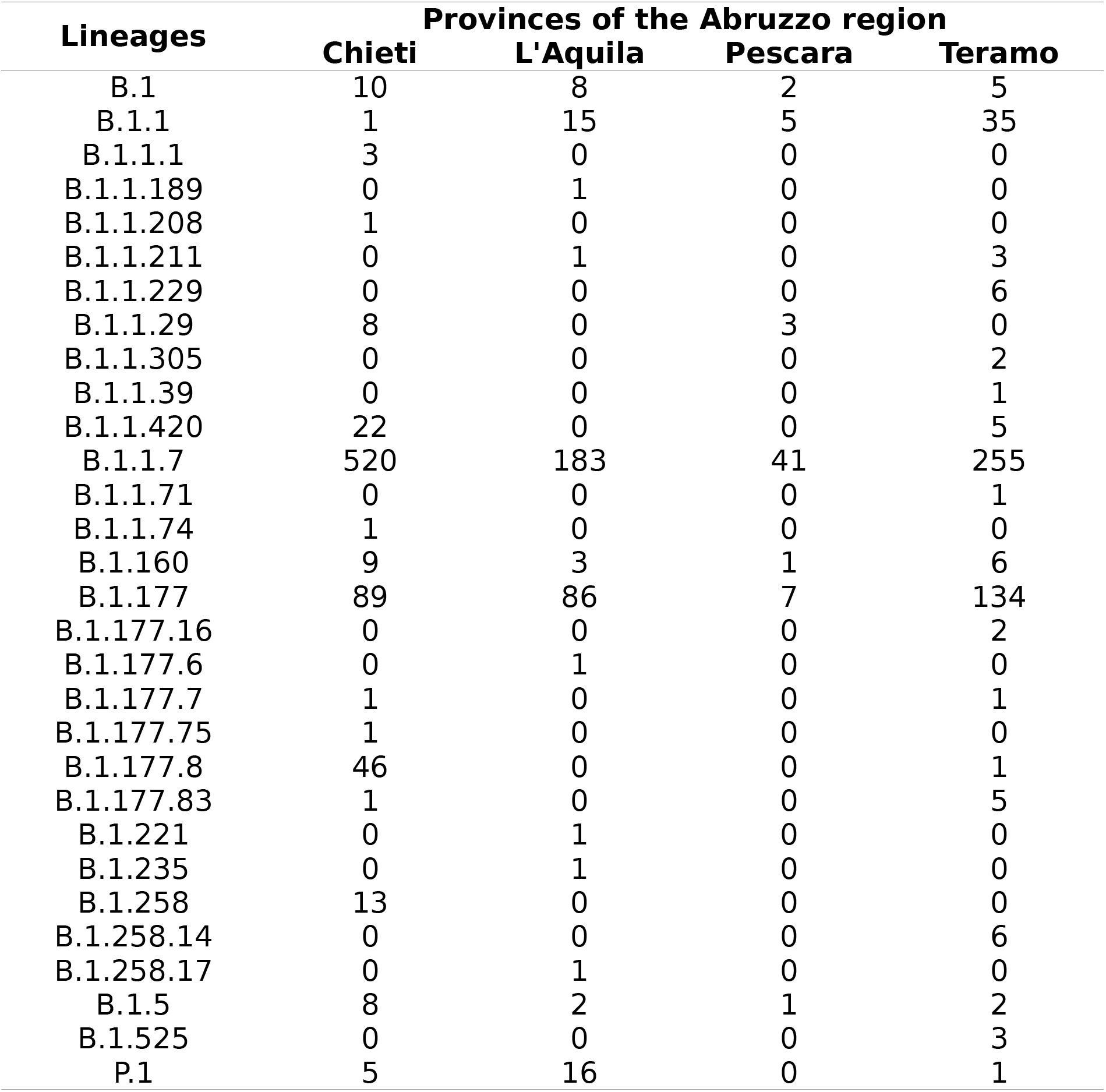
Distributions of PANGOLIN lineages from SARS-CoV-2 samples retrieved in provinces of the Abruzzo region in Italy, then shotgun sequenced and analyzed by GENPAT until April 2021 (n = 1 592).

| Lineages | Provinces of the Abruzzo region |  |  |  |
| --- | --- | --- | --- | --- |
|  | Chieti | L'Aquila | Pescara | Teramo |
| B.1 | 10 | 8 | 2 | 5 |
| B.1.1 | 1 | 15 | 5 | 35 |
| B.1.1.1 | 3 | 0 | 0 | 0 |
| B.1.1.189 | 0 | 1 | 0 | 0 |
| B.1.1.208 | 1 | 0 | 0 | 0 |
| B.1.1.211 | 0 | 1 | 0 | 3 |
| B.1.1.229 | 0 | 0 | 0 | 6 |
| B.1.1.29 | 8 | 0 | 3 | 0 |
| B.1.1.305 | 0 | 0 | 0 | 2 |
| B.1.1.39 | 0 | 0 | 0 | 1 |
| B.1.1.420 | 22 | 0 | 0 | 5 |
| B.1.1.7 | 520 | 183 | 41 | 255 |
| B.1.1.71 | 0 | 0 | 0 | 1 |
| B.1.1.74 | 1 | 0 | 0 | 0 |
| B.1.160 | 9 | 3 | 1 | 6 |
| B.1.177 | 89 | 86 | 7 | 134 |
| B.1.177.16 | 0 | 0 | 0 | 2 |
| B.1.177.6 | 0 | 1 | 0 | 0 |
| B.1.177.7 | 1 | 0 | 0 | 1 |
| B.1.177.75 | 1 | 0 | 0 | 0 |
| B.1.177.8 | 46 | 0 | 0 | 1 |
| B.1.177.83 | 1 | 0 | 0 | 5 |
| B.1.221 | 0 | 1 | 0 | 0 |
| B.1.235 | 0 | 1 | 0 | 0 |
| B.1.258 | 13 | 0 | 0 | 0 |
| B.1.258.14 | 0 | 0 | 0 | 6 |
| B.1.258.17 | 0 | 1 | 0 | 0 |
| B.1.5 | 8 | 2 | 1 | 2 |
| B.1.525 | 0 | 0 | 0 | 3 |
| P.1 | 5 | 16 | 0 | 1 |

**Figure 3:**
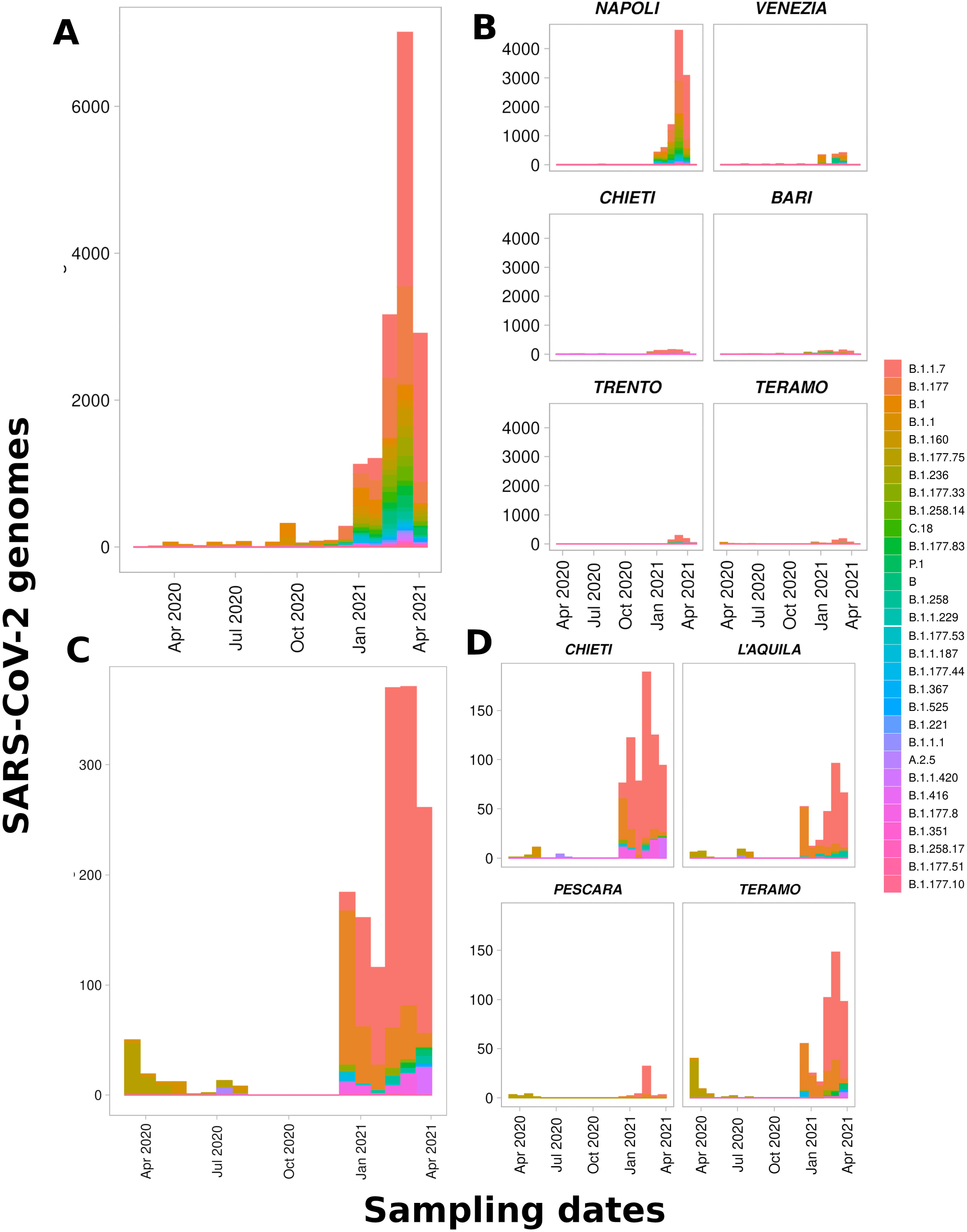
Distributions of the 30 most frequent SARS-CoV-2 PANGOLIN lineages in Italy centralized in GISAID represented at the national level (A, n = 16 529); among the 6 most frequent provinces in Italy including Napoli (n = 10 115), Venezia (n = 1 214), Chieti (n = 705), Bari (n = 661), Trento (n = 632) and Teroma (n = 501) (B, n = 13 828); among samples from the Abruzzo region mainly shotgun sequenced and analyzed in GENPAT (C, n = 1 580); and inside the provinces of the Abruzzo region including Chieti (n = 705), Teromo (n = 501), l’Aquila (n = 320) and Percara (n = 54) (D).

**Figure 4:**
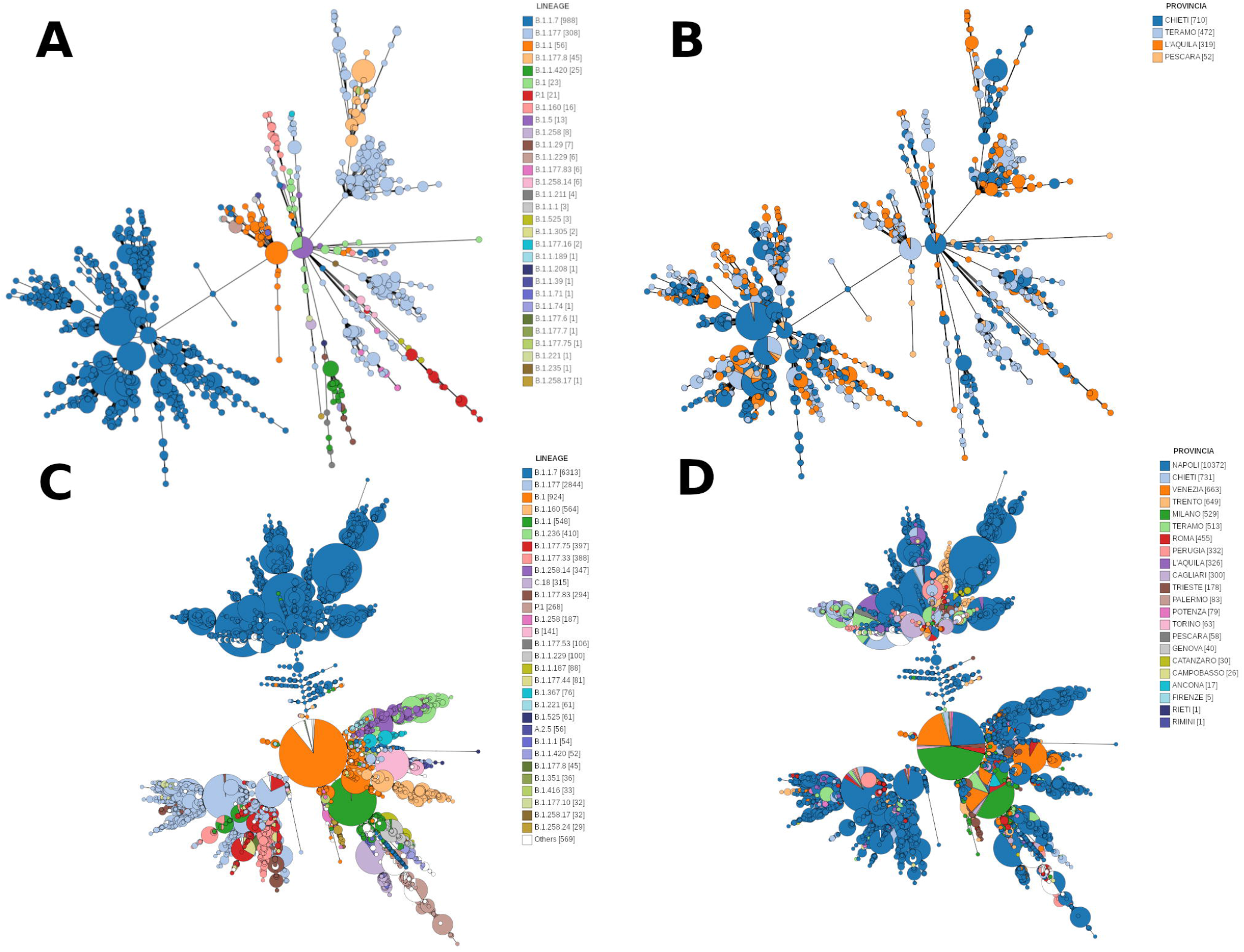
Minimum spanning tree (MST) produced by the GENPAT workflow from SARS-CoV-2 samples isolated in provinces of the Abruzzo region and sequenced by GENPAT (A and B, n = 1 553) or SARS-CoV-2 samples isolated in several Italian provinces and retrieved from GISAID (C and D, n = 15 451) with regard to lineages identified through PANGOLIN dynamic nomenclature (A and B) or Italian provinces (A and C). The effective sample sizes are shown in square brackets.

### iii/ Clustering of the MST-based method in comparison with the reference PANGOLIN lineages

In addition to the accurate identification of PANGOLIN lineages (Additional files 1 and 2), the GENPAT workflow provides MST-based clustering (Figure 1 and 2) as exemplified by trees representing SARS-CoV-2 samples isolated in the Abruzzo region and sequenced by IZSAM (Figure 4A and 4B, n = 1 553) or SARS-CoV-2 samples isolated in several Italian provinces and retrieved from GISAID (Figure 4C and 4D, n = 15 451). The effective sizes of the collections of samples (GENPAT in Additional file 1 and GISAID in Additional file 2) and MST-based clustering (GENPAT in Figure 4AB and GISAID in Figure 4CD) are slightly different at the regional (n = 1 592 *versus* n = 1 553) and national (n = 17 201 *versus* n = 15 451) levels, because these graphical representations do not include samples with missing metadata and provinces other than Abruzzo submitted to GISAID few samples isolated from this province. In view of the SARS-CoV-2 collections from GENPAT and GISAID, 30 and 176 PANGOLIN lineages were identified in the Abruzzo region and in Italy, respectively (Additional files 1 and 2). Most of the PANGOLIN lineages were grouped together by MST-based clustering (Figure 1 and 2) at the regional (Figure 4B) and national (Figure 4D) levels.

### iv/ Discrimination power of the MST-based method in comparison with the reference PANGOLIN lineages

In view of MST-based clustering and far more than the other SARS-CoV-2 lineages, the PANGOLIN lineages B.1.1.7 (n = 988 and n = 6 313) and B.1.177 (n = 308 and n = 2 844) were the mostly identified in the Abruzzo province (Figure 4B: 63% and 19%, n = 1 553) and Italy (Figure 4D: 40% and 18%, n = 15 451), respectively. Focusing on these two main PANGOLIN lineages, both lineages B.1.1.7 and B.1.177 were represented by multiple MST-based clusters (Figure 3B and 3D), emphasizing a higher discriminatory power of the MST-based method in comparison to PANGOLIN.

### v/ Speed of the MST-based method in comparison the usual phylogenomic inferences based on MSA and substitution model

While MSA and substitution model-based phylogenomic inference would require several days to several weeks to reconstruct evolution history of several hundreds of genomes with a usual computing facility (i.e. server harboring 32 Go RAM and 32 core CPUs), GENPAT estimates that 30 s and 4 s were necessary to treat 1 000 samples with the algorithms “vcf2mst.pl” and “MSTree V2”, respectively (Additional files 4 and 5).

## DISCUSSIONS

Altogether, the proper identification of lineages (i) of concern (ii), as well as the accurate (iii), discriminant (iv) and fast (v) MST-based inference, are in line with the SARS-CoV-2 surveillance needs.

### i/ Accurate GENPAT identification of PANGOLIN lineages

Due to exact match between PANGOLIN lineages identified by GENPAT and the reference GISAID (Additional files 1 and 2, n = 1 550), we recommend to identify SARS-CoV-2 lineages based on trimming, mapping, consensus building and lineage identification implemented in Trimmomatic [68], BWA [39], iVar [26] and PANGOLIN 2.0 [34], respectively (Figure 1). Faced to the diversity of methods to identify SARS-CoV-2 variants based on mapping (i.e. SAMtools [27], Freebayes [28], GATK4 [29]) and variant calling analyses (i.e. Minimap [27], Minimap2 [28], BWA [29, 30], Bowtie2 [25]), we also encourage to pursue comparisons of these methods because the SARS-CoV-2 lineages are identified based on SNPs and InDels [34], while InDels are known to be more challenging to call than SNPs [69].

### ii/ PANGOLIN lineages of concern B.1.177 and B.1.1.7 in Italy

Numerous new lineages of SARS-CoV-2 emerged since the beginning of pandemic and three of them are today considered as global variants of concern: B.1.1.7, B.1.351, and P.1 (i.e. B.1.1.248 reclassified to B.1.1.28.1) [34]. These PANGOLIN lineages B.1.1.7, B.1.351 and P1 were first detected in United Kingdom (UK) [70], South Africa [71] and Brazil [72], respectively. These lineages B.1.1.7, B.1.351, and P.1 replaced previous circulating variants in their original countries and spread to other countries in Europe, the Americas, and Asia [73]. There is an inordinate amount of concern for these three lineages because of likely reinfections due to reduced cross-protective immunity [74–76] and potential involvement in vaccine efficacy [77, 78]. While the PANGOLIN lineage B.1.177 identified frequently in Abruzzo (Table 1) and Italy (Additional file 3) corresponds to one of the first lineage identified at the beginning of the pandemic event in Italy [6], the other frequently isolated PANGOLIN lineage B.1.1.7 corresponds to the variant of concern called “UK variant” [72, 79–82]. In addition, GENPAT did not identified many samples corresponding to variants of concern named “South Africa” (B.1.351) [72, 79, 86, 87], “Japan-Brazil” (B.1.1.248 reclassified to B.1.1.28.1 -alias P.1) [72, 79, 86, 87, 89, 90], “Nigeria” (B.1.1.207) [72, 83], “Denmark” (Y453F, 69–70deltaHV) [84, 85], “UK-Nigeria” (B.1.525) [88], and “Indian” (B.1.617) [91], neither in the Abruzzo region (Figure 3C, Figure 4AB, Table 1 and Additional file 1), or Italy (Figure 3AB, Figure 4CD, Additional files 2 and 3).

### iii/ Clustering of the MST-based method in agreement with PANGOLIN

To our knowledge, it is the first time that variant calling analysis [41–43] and MST-based method [65] are combined to infer phylogenomic history of SARS-CoV-2 samples. The adaptation of the MST-based method usually used after cgMLST characterization of bacterial draft assemblies [53–57], to variant calling analysis widely used for SARS-CoV-2 investigation [27–29], allowed building of an efficient clustering workflow (Figure 1 and 2), in almost complete agreement with the outcomes of the reference PANGOLIN lineages [34]. In a near future, we plan to label the MST clusters to propose a unified method to characterize SARS-CoV-2 lineages and infer phylogenomic history at the same time.

### iv/ High discrimination power of the MST-based method

The fact that the two main PANGOLIN lineages in Italy (i.e. B.1.1.7 and B.1.177) are constituted of multiple MST clusters, emphases that the proposed method (Figure 1 and 2) presents a higher discriminatory power than PANGOLIN for SARS-CoV-2 characterization (Figure 3B and 3D). The present MST-based method (Figure 1 and 2) is also able to build MST only based on genotypes from functional annotations of variants identified in specific SARS-CoV-2 epitopes. This useful option of the algorithm “vcf2mst.pl” aims at providing graphical warnings related to SARS-CoV-2 mutations acquired in regions known to be involved in immune responses [92].

### v/ Fast minimum spanning tree from function annotations of variants

In comparison to the time consuming steps (i.e. several days or weeks for thousands samples) aiming at performing MSA from *de novo* assemblies or consensus sequences [27, 30–32], as well as phylogenomic inferences based on substitution models [27, 31, 33], the Hamming distance-based method [65] developed in the present manuscript (Figure 1 and 2) is very fast (i.e. tens of seconds to process thousands of samples). Even if this MST-based method does not root trees and does not take in account differences of evolution rates between lineages [65], this last allows fast graphical representation of SARS-CoV-2 spreading for surveillance activities requiring rapid assessment of epidemiological clusters from large amount of samples (Figure 4). According to the World Health Organization (WHO), the fast generation and global sharing of virus genomic sequences will contribute to the understanding of transmission and the design of clinical and epidemiological mitigation strategies [93]. Collaboration between public health bodies, data generators and analysts is essential to generate and use appropriately data for maximum public health benefit [93].

## CONCLUSION

The main outcomes of the present study showed that (i) GENPAT and GISAID identified identical PANGOLIN lineages, (ii) the PANGOLIN lineages B.1.177 (i.e. historical in Italy) and B.1.1.7 (i.e. “UK variant”) are major concerns today in several Italian provinces, and the new MST-based method (iii) clusters most of the PANGOLIN lineages together, (iv) with a higher dicriminatory power than PANGOLIN, (v) and faster that the usual phylogenomic methods based on MSA and substitution model. The shotgun sequencing efforts of Italian provinces, combined to a structured national management of metagenomics data, provided an accurate and fast answer supporting the system of SARS-CoV-2 surveillance in Italy. In addition, the outcomes of the present consortium involved in SARS-CoV-2 surveillance in Italy emphasized that the data sharing through GISAID is of paramount importance for supporting the international SARS-CoV-2 tracking.

## MATERIAL AND METHODS

A workflow was implemented in GENPAT during 2021 to identify SARS-CoV-2 lineages and build accurate, discriminant and fast phylogenomic inferences from several thousands of samples isolated in Italy based on shotgun metagenomics paired-end read sequencing (Figure 1).

### I/ Collections of SARS-CoV-2 samples

Two collections of SARS-CoV-2 samples were established (i.e. metadata, lineages and functional annotation of variants). The first collection includes 1 592 samples detected by IZSAM in the Abruzzo region (Italy), then shotgun sequenced and analyzed in GENPAT. Sequences were then systematically submitted by GENPAT to GISAID (https://www.gisaid.org/) [94]. The second collection includes 17 201 samples isolated from several Italian regions and retrieved from GISAID during April 2021 [94]. While samples from the first collection were treated through the whole GENPAT workflow, those from the second collection were treated through the dataset dependent part of this workflow based on information retrieved from GISAID (Figure 1).

### II/ Isolation and sequencing

Concerning the samples from the first collection, acquisition of sequencing data implied successively sampling (oropharyngeal swab transport medium or bronchoalveolar lavage), virus inactivation (PrimeStore® MTM, in BSL3 biocontainment laboratory), nucleic acid purification (MagMaxTM CORE from Thermofisher), real-time RT-PCR-based SARS-CoV-2 RNA detection (TaqManTM 2019-nCoV Assay Kit v1 or v2 from Thermofisher) [23], RNA reverse transcription through multiplexing PCR (primer scheme nCoV-2019/V1) following the ARTIC protocol (https://artic.network/) [95], cDNA purification (AMPure XP beads, Agencourt), cDNA quantification (Qubit dsDNA HS Assay Kit and Qubit fluorometer 2.0 from Thermofisher or QuantiFluor ONE dsDNA System from Promega and FLUOstar OMEGA from BMG Labtech), library preparation (Illumina DNA Prep kit) and 150 bp paired-end read sequencing (MiniSeq or NextSeq500 from Illumina).

### III/ Variant calling analysis

With the objective to avoid the time consuming MSA [44–49] and propose an accurate, discriminant and fast phylogenomic inference, the reference genome mapping [37–40], variant calling analysis [41–43] and functional annotation of variants (SNPs, MNPs and InDels) [96, 97] were preferred to *de novo* assembly [24, 36] or consensus sequences [26]. More precisly, we implemented a mapping-based variant calling analysis including functional variant annotations based on Trimmomatic [68], BWA [39], FreeBayes [42] and SNPeff [96] implemented in Snippy [98] because this workflow is fast and already well packaged in Docker (Figure 1). This read mapping and variant calling analysis were implemented with the usual SARS-CoV-2 reference genome Wuhan-Hu-1 (i.e. NC_045512).

### IV/ Identification of lineages

Faced to the rareness of other tools dedicated to lineage identification of SARS-CoV-2 (Nextstrain [99] and GISAID [100]), the lineage assignment algorithm pangoLEARN from the workflow PANGOLIN 2.0 has been implemented in the GENPAT workflow (Figure 1) to assign PANGOLIN lineages with a multinomial logistic regression-based machine learning coupled to a dynamic nomenclature of mutations associated with important functional evolution events [34]. More in details, consensus sequences were derived from BWA-based read mapping [39] with the program iVar [26] before to be used as input of the workflow PANGOLIN 2.0 [34] (Figure 1). In brief, this PANGOLIN dynamic nomenclature proposes to label major lineages with a letter starting from the earliest lineage A SARS-CoV-2 viruses closely related to the most recent common ancestor (MRCA) Wuhan/WH04/2020 (EPI_ISL_406801) isolated on 5 January 2020 from the Hubei province in China. The early representative SARS-CoV-2 sample of the lineage B was isolated on 26 December 2019: Wuhan-Hu-1 (GenBank accession no. MN908947). Then, the dynamic nomenclature assigns a numerical value for each descending lineage from either lineage A or B (e.g A.1 or B.2) following roles with corresponding criteria [34].

### V/ Phylogenomic inferences

Keeping in mind the objective to build accurate, discriminant and fast phylogenomic trees, we replaced the slow substitution model-based phylogenomic inferences [50–52] by MST inferred with the algorithm “MSTree V2” implemented in GrapeTree [65] (Figure 1). Similarly to cgMLST workflows which use alleles of orthologous genes to build MST, we propose in the present manuscript an algorithm called “vcf2mst.pl” to infer MST from functional annotation of variants (Figure 1). This algorithm “vcf2mst.pl” uses sample dependent VCF files from upstream reference genome based-variant calling analysis (Figure 1) to infer binary matrix of genotypes representing unique functional annotations of variants encoded in these VCF files (Figure 2). The three main steps of this algorithm “vcf2mst.pl” aims to (1) derive functional annotations of variants (i.e. genotypes) encoded in variant calling format (vcf) into lists of samples and genotypes (lst), (2) build a scheme of genotypes (sch) and (3) create a binary matrix of genotypes according to samples of interest (Figure 2). This algorithm “vcf2mst.pl” encodes the unique genotypes of SNPs and MNPs following the nucleotide pattern “reference genotype – position -alternative genotype” (e.g. snp:C241T), while the unique genotypes of InDels are encoded following the nucleotide pattern “position -reference genotype -alternative genotype” (e.g. ins:11287-G-GTCTGGTTTT or del:11287-GTCTGGTTTT-G). In contrast, the unique genotypes from GISAID (i.e. ZAPPO_GISAID_VCF) are encodes following the amino acid patterns “gene name _ reference amino acid _ position _ alternative amino acid” for SNPs and MNPs (e.g. NSP12_P323L or Spike_D614G), “gene name _ ins _ position _ amino acid” for insertions (e.g. NSP6_ins35VL) and “gene name _ amino acid _ position _ del” for deletions (e.g. NSP1_M85del).

## Data Availability

Metadata and consensus sequences of all the SARS-CoV-2 are available from GISAID (https://www.gisaid.org/) at the accession numbers described in supplementary information. The algorithm vcf2mst.pl is available in GitHub (https://github.com/genpat-it/vcf2mst).

## DECLARATIONS

### Ethics, consent and permissions

The results analyzed in the present study derive from the official control activities performed by the Public Health Local Authority of Abruzzo region. All human data and samples were collected ethically with consents and permissions of participants, following the “Decreto della Giunta Regionale DGR n. 194 del 2.04.2021” from the “Dipartimento Sanità della REGIONE ABRUZZO”. A local ethics committee, so-called the Ethics Committee of the National Reference Centre for Whole Genome Sequencing of microbial pathogens: database and bioinformatic analysis, ruled that no formal ethics approval was required in this particular case because the related data were openly available to the public before the initiation of the study.

## Consent for publication

Not applicable.

## Competing interests

The authors declare that they have no competing interests.

## Funding

The study was funded by the European Union’s Horizon 2020 Research and Innovation program under grant agreement No 773830: One Health European Joint Program and by the Italian Ministry of Health IZSAM 05/20 Ricerca Corrente 2020 “PanCO: epidemiologia e patogenesi dei coronavirus umani ed animali”. Mention of trade names or commercial products in this article is solely for the purpose of providing specific information and does not imply recommendation or endorsement by the IZSAM.

## Authors’ contributions

IM integrated the variant calling-based workflow in the GENPAT system with Python. ADP developed the Pearl-based algorithm “vcf2mst.pl”. NR performed the R-based graphical representation. NR drafted the manuscript and integrated comments from ADP, IM, PC, AL and CC. All authors commented and approved the final manuscript, take public responsibility for appropriate portions of the content and agree to be accountable for all aspects of the work in terms of accuracy or integrity.

## Acknowledgments

We thank especially the Italian Ministry of Health for supporting in the acquisition of high-performance computing resources.

## Abbreviations

ARDS: acute respiratory distress syndrome
CoV: coronavirus
COVID-19: coronavirus disease 19
GATK4: genomic analysis toolkit
GENPAT: Whole Genome Sequencing of microbial pathogens: data-base and bioinformatics analysis
GISAID: Global Initiative on Sharing All Influenza Data
IZSAM: Istituto Zooprofilattico Sperimentale dell’Abruzzo e del Molise Giuseppe Caporale
ML: maximum likelihood
MSA: multiple sequence alignment
MST: minimum spanning trees
NRC: National Reference Centre
RdRp: RNA-dependent RNA polymerases
SARSr-CoVs: SARS-related coronaviruses
SARS-rCoV: severe acute respiratory syndrome-related virus
(UK): United Kingdom
WHO: World Health Organization

## Availability of data and materials

Metadata and consensus sequences of all the SARS-CoV-2 are available from GISAID (https://www.gisaid.org/) at the accession numbers described in supplementary information. The algorithm “vcf2mst.pl” is available in GitHub (https://github.com/genpat-it/vcf2mst).

## SUPPLEMENTARY INFORMATION

**Additional file 1:** Metadata and PANGOLIN lineages of the dataset of SARS-CoV-2 samples isolated by IZSAM in provinces of the Abruzzo region (Italy), then shotgun sequenced and analyzed in GENPAT until April 2021 (n = 1 592).

**Additional file 2:** Metadata and PANGOLIN lineages of the dataset of SARS-CoV-2 samples isolated from several Italian provinces and retrieved by GENPAT from GISAID until April 2021 (n = 17 201).

**Additional file 3:** Distributions per Italian provinces (n = 25) of the SARS-CoV-2 PANGOLIN lineages in Italy (n = 176) retrieved by GENPAT from GISAID until April 2021 (n = 17 201).

**Additional file 4:** Newick file inferred through variant calling-based analysis, binary matrix of functional annotations of variants (SNPs, MNPs and InDels) with the program “vcf2mst.pl” and Hamming-like distance-based minimum spanning tree (MST) implemented in GrapeTree (“MSTree V2”), from the dataset of SARS-CoV-2 samples isolated by IZSAM in provinces of the Abruzzo region (Italy), then shotgun sequenced and analyzed in GENPAT until April 2021 (n = 1 553).

**Additional file 5:** Newick file inferred through variant calling-based analysis, binary matrix of functional annotations of variants (SNPs, MNPs and InDels) with the program “vcf2mst.pl” and Hamming-like distance-based minimum spanning tree (MST) implemented in GrapeTree (“MSTree V2”), from the dataset of SARS-CoV-2 samples isolated from several Italian provinces and retrieved by GENPAT from GISAID until April 2021 (n = 15 451).

## Notes

### Competing Interest Statement

The authors have declared no competing interest.

### Clinical Trial

The present study is neither a clinical trial, nor a prospective studies.

### Funding Statement

The study was funded by the European Union Horizon 2020 Research and Innovation program under grant agreement No 773830: One Health European Joint Program and by the Italian Ministry of Health IZSAM 05/20 Ricerca Corrente 2020 PanCO: epidemiologia e patogenesi dei coronavirus umani ed animali. Mention of trade names or commercial products in this article is solely for the purpose of providing specific information and does not imply recommendation or endorsement by the IZSAM.

### Author Declarations

The results analyzed in the present study derive from the official control activities performed by the Public Health Local Authority of Abruzzo region. All human data and samples were collected ethically with consents and permissions of participants, following the Decreto della Giunta Regionale DGR n. 194 del 2.04.2021 from the Dipartimento Sanità della REGIONE ABRUZZO. A local ethics committee, so‑called the Ethics Committee of the National Reference Centre for Whole Genome Sequencing of microbial pathogens: database and bioinformatic analysis, ruled that no formal ethics approval was required in this particular case because the related data were openly available to the public before the initiation of the study.

